# Variation in Haemostasis and VTE Prophylaxis in Elective Adult Cranial Neurosurgery: A Global Survey of Perioperative Practice

**DOI:** 10.64898/2026.04.14.26350905

**Authors:** AS Pandit, T Chaudri, Z Chaudri, AM Vasilica, J Dhaliwal, Z Sayar, H Cohen, JP Westwood, AK Toma

## Abstract

**Background:** Venous thromboembolism (VTE) remains a major cause of perioperative morbidity in cranial neurosurgery, yet clinical practice varies widely, and formal guidelines are inconsistent. Understanding internationally sampled neurosurgical practice is essential for informing consensus and future trials.

**Methods:** An international, 2-stage cross-sectional, internet-based survey was conducted. Practising neurosurgeons performing elective adult cranial surgery were eligible. Descriptive statistics were used to summarise practice. Responses covered patterns of pre-operative haemostasis decision making, use and timing of mechanical and/or chemical prophylaxis, use of perioperative imaging prior to anticoagulation, and frequency of clinical assessment for VTE. Associations with geographical income status, subspecialty, and years post-certification were statistically tested. Practice heterogeneity was quantified and contextual influence was summarised using mean effect sizes across stratifying variables in order to determine domains of true equipoise.

**Results:** Of 585 responses, 456 (78%) met criteria for inclusion: representing 322 units across 78 countries (71% high-income). Thirteen per cent reported no departmental VTE plan; 23% followed no guidelines and 12% used multiple. Routine pre-operative testing almost universally included haemoglobin/platelets/haematocrit, with fibrinogen more common in high-income settings. Compared with high-income country respondents, low- and middle-income respondents reported higher haemoglobin transfusion thresholds (>90 g/dL; p<0.001) and shorter antiplatelet interruption (p≤0.03), and less frequent outpatient VTE assessment (p<0.001). Mechanical prophylaxis was common (TEDs 81%, IPC 62%), typically started pre-or intra-operatively. Among those completing the chemoprophylaxis section (n=310), 57% required a CT or MRI scan before LMWH which was then initiated on average 31.4 hours after surgery. 1% of respondents did not routinely use LMWH. Many clinical decisions demonstrated statistical equipoise ie. high heterogeneity with low contextual influence.

**Conclusion:** Peri-operative haemostasis and VTE prophylaxis practices in adult elective cranial neurosurgery vary substantially worldwide, with some decisions reflecting geographical or socioeconomic differences and many others reflecting true clinical equipoise rather than contextual determinants. By mapping contemporary real-world practice across diverse health-system contexts, this study provides a necessary empirical foundation for rational trial design and future guideline development.

## Introduction

The management of venous thromboembolism (VTE) prophylaxis in cranial neurosurgery is defined by a narrow therapeutic window between thromboprophylaxis and the prevention of catastrophic intracranial hemorrhage (ICH). The neurosurgical population is inherently predisposed to VTE, driven by a multifactorial risk profile that includes extended operative duration, limited mobility, and often, the hypercoagulability associated with malignancy and steroid exposure. Unlike many types of extracranial surgery, even minor postoperative bleeding in confined intracranial compartments can result in neurological deterioration, need for reoperation, permanent disability and death.

Reported rates of postoperative VTE following cranial neurosurgery vary widely in the literature, reflecting differences in case mix, surveillance strategies, and prophylaxis practices. Symptomatic VTE has been reported in approximately 1–5% of patients and total VTE rates rise substantially when routine screening is employed, with some series suggesting incidences exceeding 20% [1,2]. These thromboembolic events are associated with significant morbidity, prolonged hospitalisation, and increased healthcare utilisation, and remain a leading cause of preventable postoperative mortality in surgical populations [3,4]. Conversely, postoperative ICH rates range from approximately 1–14%, but again are dependent on the type of surgery and surveillance and prophylaxis strategies, and whether the hematoma warranted a return to theatre [5–8]. The fear of precipitating intracranial bleeding for many remains a major driver of conservative or delayed pharmacological VTE prophylaxis in neurosurgical practice [9].

Clinical decision-making is further complicated by the intrinsically multifactorial nature of coagulation risk in cranial neurosurgery. Patient-specific factors (age, comorbidity, malignancy, prior VTE), disease-related factors (tumour type, inflammatory state), intra-operative variables (operative duration, blood loss, use of haemostatic adjuncts), and postoperative considerations (drains, reoperation risk, neurological status) could interact dynamically across the peri-operative period [10,11]. This complexity makes uniform, protocol-driven approaches challenging and likely contributes to substantial heterogeneity in real-world practice, both within and between healthcare systems.

The evidence base guiding the choice, timing, and duration of chemical VTE prophylaxis in cranial neurosurgery remains limited. High-quality randomised controlled trials are scarce, many studies are underpowered for bleeding outcomes, and existing data are frequently extrapolated from heterogeneous surgical populations [12]. As a result, international guidelines—including those from the United Kingdom’s National Institute for Health and Care Excellence (NICE) [13], the American Society of Hematology (ASH) [14], the Australia & New Zealand Working Party on the Management and Prevention of Venous Thromboembolism [15] and the Thrombosis and Haemostasis Society of Australia and New Zealand (THANZ) [16], European Society of Anaesthesiology and Intensive Care (ESAIC) [17] — provide cautious and, at times, absent or divergent recommendations, particularly with respect to pre-operative cessation of antiplatelet agents and postoperative initiation timing and duration of low-molecular-weight heparin (Table 1).

**Table 1.** Summary and synthesis of relevant regional, national and international guideline recommendations for peri-operative haemostasis and VTE prophylaxis specific to adult elective cranial neurosurgery. ASH, American Society of Haematology; IPCs, intermittent pneumatic compression stockings; LMWH, low-molecular weight heparin; NICE, National Institute for Health and Care Excellence; TEDs, thromboembolism deterrent stockings; TE, thrombo-embolic; and UFH, unfractionated heparin.

Importantly, implementation of these recommendations varies considerably across neurosurgical centres. Resource availability, institutional infrastructure, access to imaging, pharmacological agents, laboratory testing, and critical care support differ substantially geographically between high-income countries (HICs) and low- and middle-income countries (LMICs) [18,19]. These systemic factors may influence risk tolerance, thresholds for pharmacological prophylaxis, timing of initiation, and management of bleeding complications. In LMIC settings in particular, limited access to rapid imaging, blood products, or re-operation capacity may favour more conservative approaches [18,19], whereas differing VTE surveillance practices may also influence perceived thromboembolic risk [2].

In this context, systematic evaluation of contemporary VTE prophylaxis practice is essential. Characterising current patterns of care enables quantification of practice heterogeneity, assessment of how peri-operative decision-making varies across healthcare systems and income settings and identification of areas of genuine clinical equipoise. Such data are critical to the design of future pragmatic randomised controlled trials, ensuring that eligibility criteria, intervention strategies, timing of prophylaxis, and outcome measures are grounded in real-world practice and are generalisable across diverse clinical environments [20].

The primary aim of this international survey was to characterise contemporary peri-operative haemostasis and VTE prophylaxis practices in adult elective cranial neurosurgery. Specifically, we sought to evaluate heterogeneity in pre-operative and post-operative decision-making, including approaches to risk stratification and the timing and use of mechanical and pharmacological prophylaxis; and to examine variation associated with clinician experience, geographical region, and national income setting. The secondary aim was to quantify the extent and drivers of this variation in order to distinguish true clinical equipoise from context-dependent practice differences. By delineating areas of uncertainty and consensus, this study aims to inform future guideline development and support the rational design of prospective interventional studies in neurosurgical VTE prevention.

## Methods

### Study design and ethical approval

This was a global, cross-sectional study conducted using a structured, internet-based survey. The survey was designed and reported in accordance with the Checklist for Reporting Results of Internet E-Surveys (CHERRIES). Ethical approval was obtained through a minimal risk pathway and granted exemption by the University College London (UCL) Research Ethics Committee. Extracted data contained no personal identifiers or institutional names, and all data were anonymised and analysed in aggregate.

### Survey development

A multidisciplinary team comprising consultant neurosurgeons, haematologists, perioperative care specialists and academics with experience of surgical survey design, first designed the survey to capture current practice regarding haemostasis and VTE prophylaxis in elective adult cranial neurosurgery. The instrument was developed through an iterative consensus process, piloted for clarity and usability, and reviewed for face validity by neurosurgical peers. The final version included closed-ended multiple-choice questions and free-text fields, covering the following domains related to: (i) pre-operative haemostasis decision-making (including cessation of blood thinners); (ii) post-operative mechanical and chemical VTE prophylaxis; (iii) VTE surveillance and initial management; (iv) self-reported rates of VTE and ICH and use of guidelines. The number of questions were counterbalanced against the total survey time which was limited to less than 10 minutes.

### Survey distribution and management

The survey was disseminated internationally between November 2023 and May 2025 via multiple channels: (i) neurosurgical mailing lists (including via national, European and international societies) and newsletters; (ii) social media including X, LinkedIn, and ResearchGate and (iii) personal and institutional academic networks. Informed consent was gained by respondents agreeing to proceed with the survey after the introduction window. Participation was voluntary and open to practising neurosurgeons involved in cranial surgery. Multiple reminders were issued to encourage completion.

Anonymised responses were collected using a secure, GDPR-compliant online survey platform (Qualtrics). Built-in logic controls enabled tailored question pathways based on prior answers, improving relevance and data quality. Respondents who were not actively performing cranial neurosurgery were excluded.

### Data Analysis

Quantitative data were analysed using descriptive and summary statistics (e.g. counts, proportions, means, medians). Where possible prophylaxis practices were stratified by subspecialty (e.g., neuro-oncology, vascular, skull base), years of post-board certification surgeon experience and by geographical region (Africa, Asia, Europe, Americas, Australasia), and World Bank (country) income status. Holm-Bonferroni correction was used in cases for multiple comparisons.

To determine priorities clinical equipoise (i.e. unexplained variation), we employed a dual-metric framework. Practice heterogeneity was quantified using Shannon Entropy for categorical variables and the Coefficient of Variation for quantitative thresholds and then normalised. To assess the extent to which this variation was driven by professional or demographic determinants, we calculated an influence score for each domain, averaging effect sizes derived from Cramer’s V, Eta-squared, Epsilon-squared, and Spearman’s Rho across stratified variables: income group, continent, subspecialty, and years of experience. Clinical domains characterized by high heterogeneity (***H*** > 0.50) and low mean influence (***I*** < 0.20) were identified as areas of “True Equipoise” [26] and primary candidates for trial consideration, while domains with high influence scores were classified as reflecting contextual variation.

## Results

### Respondent characteristics

Of 585 total responses, 456 met the criteria for inclusion (i.e. were neurosurgeons with at least some adult elective cranial practice and who completed at least 90% of the survey questions) and were included in the final analysis (Table 2). These data represent 322 neurosurgical units across 78 countries, covering approximately 48% of nations with at least one functioning neurosurgical center (Figure 1). While Europe accounted for many respondents (60%), there was significant representation from the Americas (18%), Asia (15%), Africa (6%), and Australasia (1%). By economic classification, 325 (71%) responses originated from High-Income Countries (HIC) and 131 (29%) from Low- and Middle-Income Countries (LMIC). Significant collinearity was observed (Cramer’s V=0.60) between respondent geography and income status—most notably within the European cohort, which was 91% High-Income. Consequently, country income status rather than geographical location was prioritised as a primary determinant for subsequent analysis.

**Figure 1.**
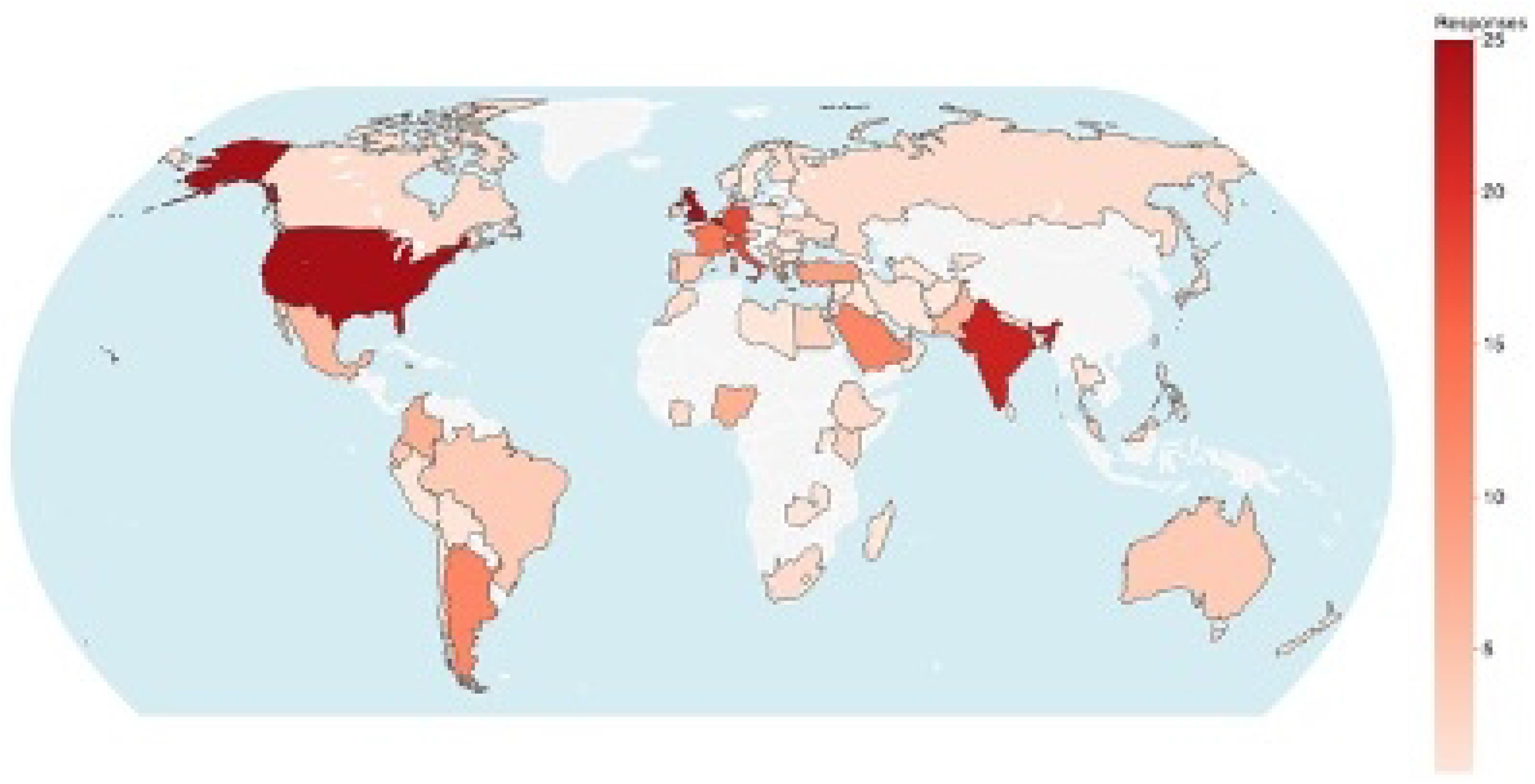
Chlorpleth map showing frequency of respondents by country. Higher frequency responses are shown as darker red. Countries with no responses were made blank

**Table 2.** Respondent demographics. *for survey efficiency, respondents were asked to declare only one subspeciality. ** includes anterior skull base and pituitary. **✝** ‘Other’ respondents had a paediatric, spine, trauma or peripheral nerves primary practice with some elective adult cranial activity

Among respondents who declared a primary subspecialty, 86% reported a cranial-focused practice (neuro-oncology, skull base, vascular, functional, pituitary, epilepsy, or adult hydrocephalus), while 14% selected “other”, most commonly spine- or trauma-dominant practice, but with some elective adult cranial activity.

A formal response rate could not be calculated because the survey used open, multichannel dissemination rather than a closed denominator-based sampling strategy.

### Guidelines and local VTE prophylaxis plans

Thirteen percent of respondents (56) stated there was no formal VTE plan in the department. Respondents were asked which national or international guideline they themselves utilised, if any (Table 1, Figure 2A). 23% of respondents (105) declared that they did not follow any specific guideline while 12% (53) stated they employed multiple guidelines for their practice. Most participants reported using guidelines specific to their geographical catchment where available; however, across all continents a substantial minority relied on guidance developed outside their own healthcare setting, most commonly those from ASH, NICE, and European societies (Figure 2B).

**Figure 2.**
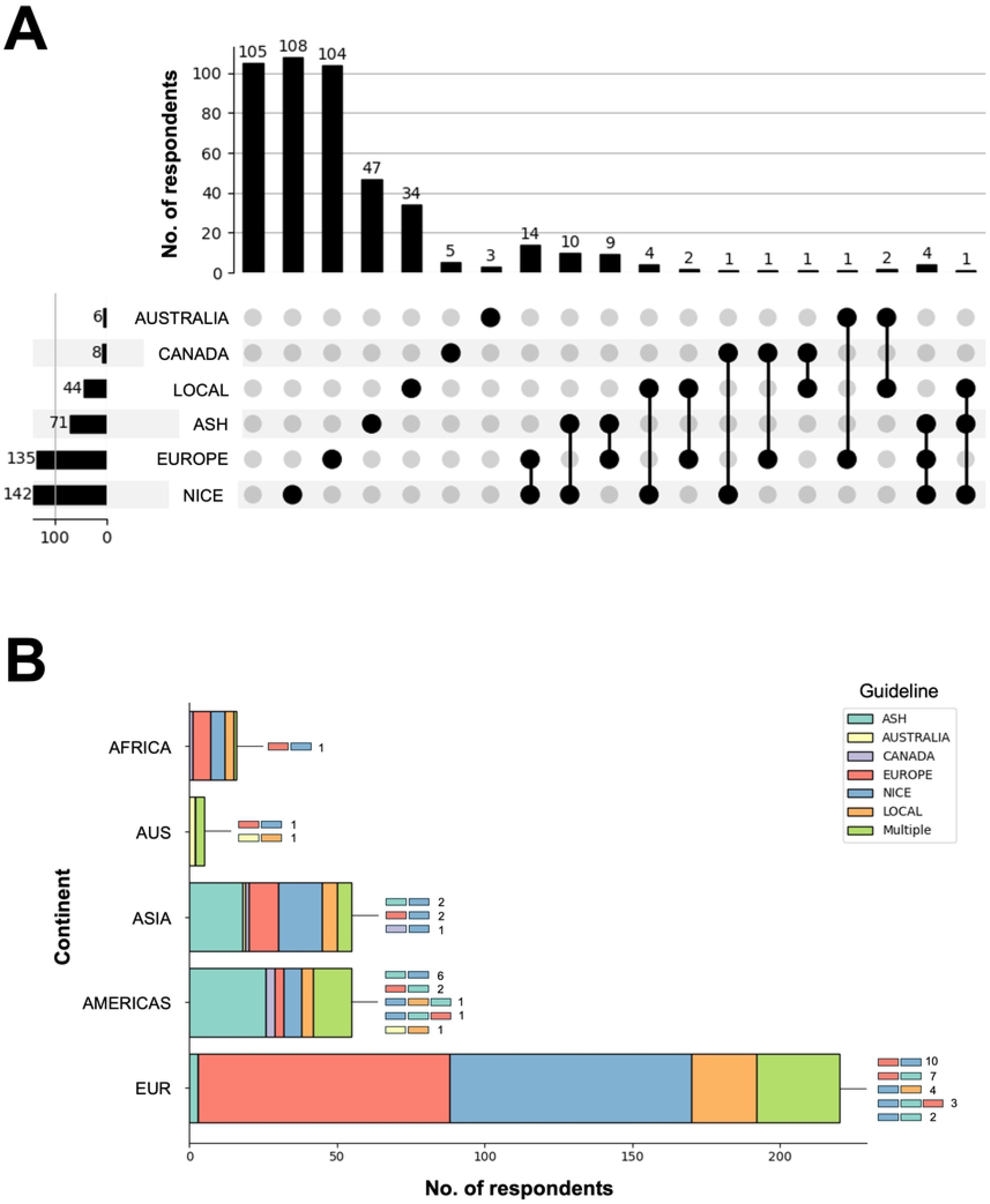
(A) UpSet plot and (B) stacked bar chart showing usage of national / international VTE guidelines. AUS = Australasia, AUSTRALIA = Australia National Health and Medical Research Council / Australia and New Zealand Working Party on the Management and Prevention of Venous Thromboembolism; CANADA = Thrombosis Canada; LOCAL = other local regional or national guidelines; ASH = American Society of Hematology; EUR = Europe, EUROPE = European Society of Anaesthesiology and Intensive Care; NICE = National Institute of Clinical Excellence.

### Decision-Making for Pre-operative Haemostasis

The majority of respondents (96%) had access to a haematology specialist or an anaesthetist for specific patients with clotting or bleeding disorders. The most frequent reasons for referral were patients with a coagulopathy (85%) or an abnormal coagulation screen or blood film (77%), or having a pro-thrombotic condition (75%). Less frequent were referrals for patients requiring a peri-operative blood transfusion and those on various types of anticoagulants or antiplatelets (Figure 3A). Respondents from LMIC countries were significantly more likely to cite DOAC (χ^2^ = 8.9, p = 0.003), antiplatelet (χ^2^ = 27.7, p < 0.0001) and warfarin usage (X^2^ = 50.7, p < 0.0001) as reasons for referral and less likely to cite blood transfusion (χ^2^ = 6.7, p = 0.01).

**Figure 3.**
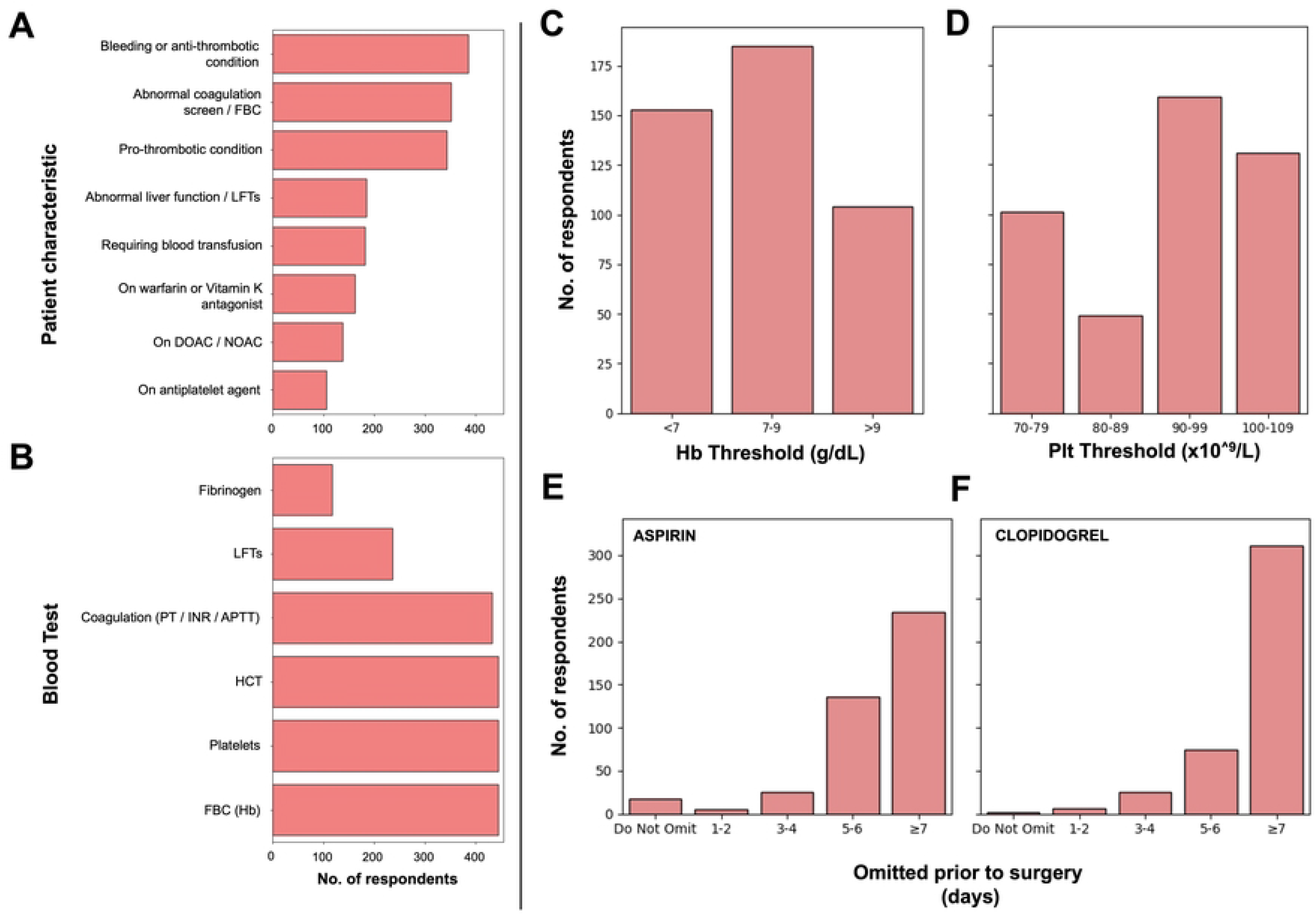
Pre-operative haemostasis and decision making. (A) Reasons for referral to a haematology specialist or an anaesthetist for specific patients with clotting or bleeding disorders. (B) Routine pre-operative blood tests. (C) Haemoglobin (Hb) and (D) Platelet (Plt) thresholds for pre-operative transfusion. Number of days an antiplatelet agent was omitted prior to surgery for (E) Aspirin and (F) Clopidogrel.

The most common routine set of blood tests taken prior to an elective cranial neurosurgical operation comprised haemoglobin, platelet count, and haematocrit for over 95% of respondents, whereas liver function tests (52%) and fibrinogen (26%) were less requested (Figure 3B). The latter was found to be significantly more requested by HI countries (χ^2^ = 4.8, p =0.03).

Seventy-four per cent of respondents reported that they would consider pre-operative red blood cell transfusion at haemoglobin thresholds of 70–90 g/L or lower (Figure 3C). This practice was not significantly associated with respondent seniority or subspecialty interest. In contrast, respondents practising in LMICs demonstrated a greater propensity to transfuse at higher haemoglobin thresholds (>90 g/L; χ^2^ = 12.3, p < 0.001). Platelet transfusion thresholds exhibited greater variability overall, although the range of 90–99 × 10^9^/L was most frequently used (Figure 3D).

Despite most respondents stopping antiplatelet agents ≥7 days pre-operatively, nearly half discontinued aspirin and over a quarter stopped clopidogrel for less than 6 days or not at all (Figure 3E,F). High-income countries were found to omit antiplatelets for longer pre-operatively (Aspirin: HIC mean = 6.3 days, LMIC mean = 6.0 days, T = 2.2, p = 0.03; Clopidogrel: HIC mean = 6.6 days, LMIC 6.1 days, T = 3.3, p = 0.001).

### Decision-making for mechanical VTE prophylaxis

81% of respondents used anti-embolism stockings (TEDS) peri-operatively, and 62% used intermittent pneumatic compression (IPC) devices. Of those who endorsed their use, the majority opted to initiate either before or during the surgical procedure (TEDS: 89%, IPCs: 88%), rather than afterward. This did not vary statistically with geography, subspeciality or seniority.

### Decision-making for chemical VTE prophylaxis

Questions relating to this domain were restricted to respondents who had identified a primary adult elective cranial subspecialty. Consequently, complete responses for this section were only available from 310 participants (68%). Of these, 57% stated they would request a CT or MRI scan prior to starting low molecular weight heparin - at a mean time of 20.4 hours post-surgery (SD=9.8). Very few participants (n=4, 1%) stated that they would *not* routinely give LMWH after surgery. When given, LMWH was started at a mean of 31.4 hours (SD = 14.9, median = 24, IQR = 24-48) after surgery. This did not appear to significantly vary by seniority, geographical income or subspeciality following multiple comparison correction.

All participants were asked which factors, if any, would prompt them to initiate chemoprophylaxis earlier than routine. These factors related to the patient, any underlying haematological or medical conditions and agents used intraoperatively (Figure 4). 399 patients (88%) provided responses for this section. Age was more frequently reported as considerations for earlier chemoprophylaxis in LMIC settings (χ^2^ = 24.1, *p* < 0.0001). Subspeciality differences were also found with, for example, skull-base and neuro-oncology placing a significantly greater emphasis on raised BMI as a factor for earlier prevention, as compared to functional.

**Figure 4.**
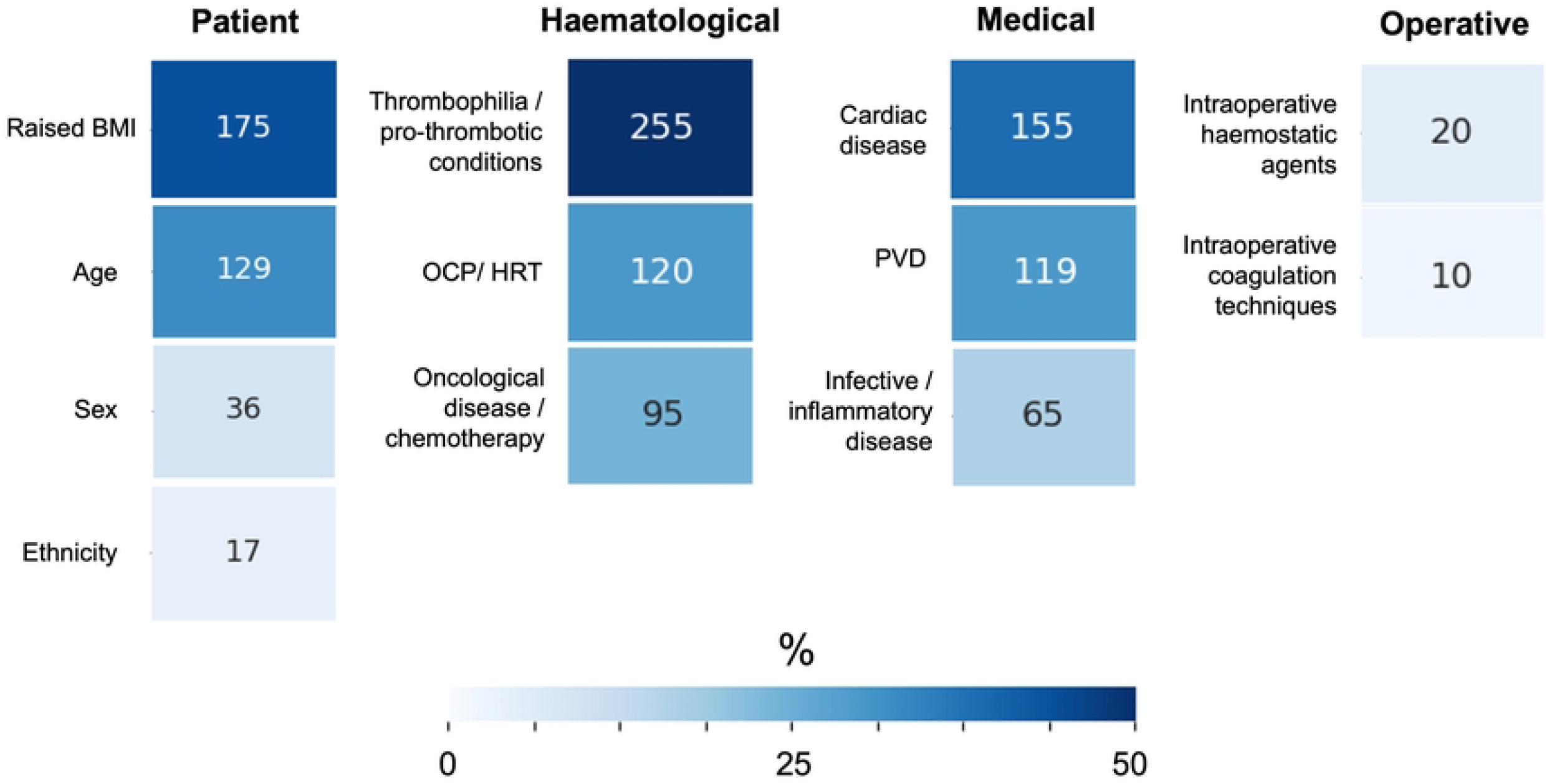
Factors prompting earlier VTE chemoprophylaxis. BMI = body mass index, HRT = hormone replacement therapy, OCP = oral contraceptive pill, PVD = peripheral vascular disease. Numbers of respondents are given in each box, while the color corresponds to the % of the available cohort.

**Figure 5.**
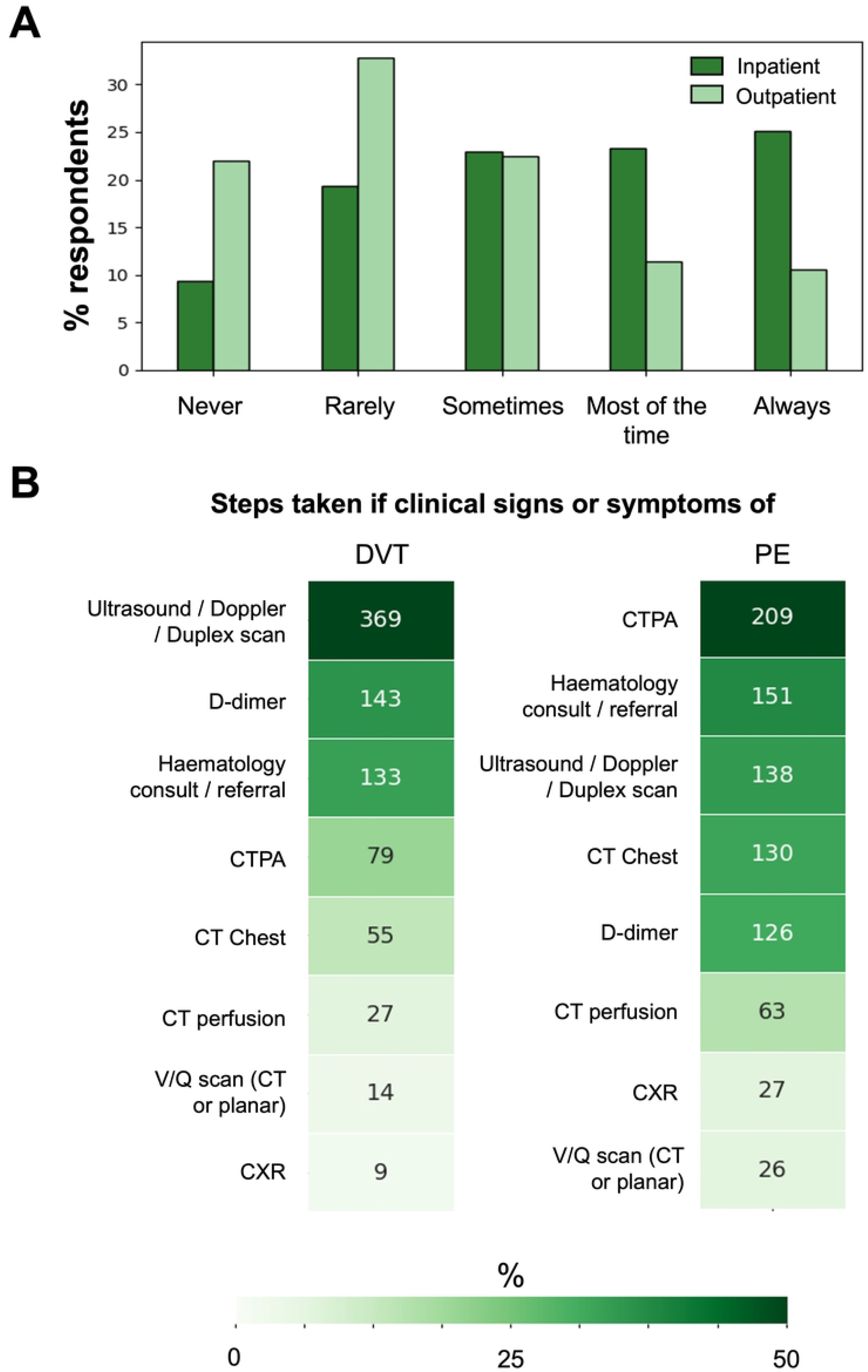
VTE surveillance and management. (A) Percentage of respondents who assessed for clinical signs and symptoms of VTE in inpatient and outpatient settings. (B)Steps taken if clinical signs or symptoms of a deep vein thrombosis (DVT) and pulmonary embolism (PE). CTPA = CT Pulmonary Angiogram, V/Q = ventilation/perfusion

### VTE surveillance and management

Respondents were asked how often they would assess clinical signs or symptoms of VTE. This was reported more frequently in the inpatient than outpatient practice, with most respondents indicating that inpatient VTE assessment was performed most of the time or always, whereas outpatient assessment was less consistent and more widely distributed across response categories. Respondents practising in HI countries reported less frequent outpatient VTE assessment compared with those practising in LMIC settings (χ^2^ = 21.5, p < 0.001).

Most respondents elected to investigate deep venous thrombosis (DVT) with an ultrasound, doppler or duplex scan (92%), and a pulmonary embolism (PE) with a CT pulmonary angiogram (52%) as shown in Figure Yet, there was greater divergence in choosing other investigation modalities and only around a third chose to consult with their local haematology team if a VTE occurred.

### Heterogeneity, influence and clinical equipoise

Across several peri-operative domains, many clinical decisions were characterised by high normalised heterogeneity coupled with low mean influence, consistent with a state of true clinical equipoise in which consensus was lacking (Table 3). In contrast, selected decisions within the pre-operative and late post-operative phases—most notably referral to haematology and investigation for suspected DVT or PE—demonstrated the highest mean effect sizes (ranging from 0.280 to 0.319). These practices were, however, strongly shaped by contextual determinants, particularly geography or income setting and clinician experience. Conversely, intra-operative and early post-operative practices, including timing of post-operative imaging, and factors for earlier chemoprophylaxis exhibited high heterogeneity alongside overall low mean influence, suggesting a pervasive absence of standardised practice rather than context-driven variation.

**Table 3.** Heterogeneity, influence and clinical equipoise of study variables of interest. If normalised heterogeneity was high > 0.5, and mean effect size was low < 0.2, true equipoise was present (highlighted in grey). Hb = haemoglobin; IPC = intermittent pneumatic compression, TED = thrombo-embolism deterrent.

## Discussion

### Summary of key findings

In this large international survey of adult elective cranial neurosurgery, spanning 322 units across 78 countries, we identified substantial and persistent heterogeneity in peri-operative haemostasis and VTE prophylaxis. While access to specialist haematology support and the use of mechanical prophylaxis were near-universal, marked variation was observed in pre-operative haemostasis strategies, timing of post-operative imaging and chemoprophylaxis initiation, approaches to VTE surveillance, as well as reliance on local, national, or international guidelines.

Certain practice patterns were associated with systemic contextual factors. For instance, LMICs favoured more conservative decision making with shorter pre-operative antiplatelet interruption, higher haemoglobin thresholds for red blood cell transfusion, and less extensive routine blood testing, while post-operatively they reported earlier chemoprophylaxis for selected patient demographics and more frequent VTE clinical surveillance compared with high-income settings. Clinician-level factors also influenced decision-making; greater experience was associated with higher utilisation of mechanical prophylaxis, and subspecialty differences were most apparent in post-operative risk stratification, with specific subgroups placing greater emphasis on earlier chemoprophylaxis.

Despite these associations, quantitative heterogeneity analysis demonstrated that many core decisions were characterised by high heterogeneity with low overall contextual influence, consistent with clinical equipoise. Taken together, these findings suggest that much of the observed global variation reflects unresolved uncertainty rather than structural constraint, underscoring the need for harmonised guidance and the prioritisation of pragmatic, internationally generalisable trials in neurosurgical VTE prevention.

### Interpretation and context

Venous thromboembolism remains one of the leading preventable causes of post-operative morbidity and mortality in neurosurgery ([27]). Despite advances in surgical technique, anaesthesia, and peri-operative care, our findings demonstrate that VTE prophylaxis practices continue to vary widely among neurosurgeons worldwide. This heterogeneity is likely multifactorial and reflects the intrinsic tension between thromboembolic prevention and the potentially catastrophic consequences of intracranial haemorrhage. Although international guidelines exist, they offer cautious and, at times, divergent recommendations—particularly regarding the timing and prerequisites for initiating chemoprophylaxis—resulting in substantial reliance on individual clinician judgement or regional custom rather than uniform standards [28]. This is underscored by the observation that nearly one-quarter of respondents reported following no formal guideline, whether local or from a major professional society, consistent with prior reports of fragmented governance in neurosurgical VTE prevention [9,29,30].

Our results corroborate and extend findings from earlier national and international surveys of neurosurgical VTE prophylaxis practice. Earlier work from the United Kingdom, Canada, and the United States consistently demonstrated marked inter-unit and inter-clinician variation, particularly in the use and timing of chemical prophylaxis [9,29,30]. In line with these studies, we observed near-universal use of mechanical prophylaxis, with increasing uptake among more experienced surgeons. However, our survey also demonstrates a near-universal intention to use post-operative chemoprophylaxis following elective cranial surgery, with very few respondents reporting a purely mechanical-only strategy. This represents a notable shift compared with earlier US-based surveys, where a substantial minority of surgeons avoided anticoagulation altogether after uncomplicated tumour craniotomy.

Our data further demonstrate that some elements of practice variation are systematically associated with contextual and clinician-level factors. Respondents practising in LMICs tended to favour more conservative peri-operative strategies, including shorter antiplatelet omission, higher haemoglobin thresholds for transfusion, less extensive routine laboratory testing, and greater post-operative VTE clinical surveillance. These patterns are consistent with prior global surgery literature suggesting that LMIC settings may adopt risk-averse strategies in the face of limited access to post-operative critical care, advanced imaging, blood products, or rapid rescue interventions for haemorrhagic complications [18]. Higher transfusion thresholds and earlier chemoprophylaxis for selected patient demographics may similarly reflect reduced tolerance for physiological reserve loss and the practical challenges of delayed recognition or treatment of thrombotic events in resource-constrained environments.

Subspecialty and experience also influenced decision-making. Greater post-certification experience was associated with increased use of mechanical prophylaxis, suggesting a learned preference for low-risk preventive strategies. Subspecialty differences were most evident in post-operative risk stratification, with skull-base and neuro-oncology surgeons placing greater emphasis on earlier chemoprophylaxis for certain patient factors, whereas functional specialists were comparatively less influenced by such factors. These findings align with prior observations that disease-specific risk perception—such as haemorrhagic risk in intracranial tumour surgery—shapes prophylaxis strategies. This implies that a single approach may not be universally applicable across all neurosurgical contexts [9,31].

Although several practices were statistically associated with selected covariates: income setting, subspecialty, and clinician experience, these factors accounted for only a small proportion of overall practice variation. Quantitative heterogeneity analysis demonstrated that many core decisions were characterised by high heterogeneity with low contextual influence, indicating that variation persists even within comparable settings and supporting the presence of genuine clinical equipoise rather than context-driven divergence. While previous neurosurgical surveys have described similar variability, our findings are the first to formally quantify this dissociation.

### Limitations and strengths

Several limitations must be acknowledged. Responses were self-reported and may reflect intended rather than actual practice. Participation was uneven across regions, with some over-representation from high-income countries and variable country-level response numbers, limiting the interpretation of national practices. Some responses were incomplete and excluded from analysis, and the questionnaire did not capture all potentially relevant factors, including specific LMWH regimens, institutional protocols, or case-mix. The prolonged survey period facilitated broad international participation but introduces the possibility that individual practices evolved during data collection. Finally, the absence of patient-level outcome data precludes assessment of the comparative effectiveness or safety of different prophylaxis strategies. While ICH and VTE rates were provided, it was felt beyond the scope of this study to perform further predictive modelling to assess key contributory variables.

Despite these limitations, this study has several notable strengths. It represents, to our knowledge, the first large-scale international survey focused specifically on peri-operative haemostasis and VTE prophylaxis in adult elective cranial neurosurgery. The inclusion of respondents across subspecialties, experience levels, and economic settings provides a broad and contemporary international perspective. A formal response rate could not be calculated due to the open, international dissemination strategy. However, the sampled cohort spanned 322 units and, within the analysable dataset, the median number of responses per unit was. We therefore interpret these findings as an international sample of reported practice rather than a definitive census of global neurosurgical behaviour. In some regions, the coverage was, including near-complete unit-level participation in the United Kingdom. Importantly, the analytical approach moved beyond descriptive reporting by quantifying heterogeneity and contextual influence, allowing identification of areas of true clinical equipoise and providing a strong foundation for future guideline development and pragmatic trial design.

### Conclusion

This study demonstrates substantial global variation in peri-operative VTE prophylaxis in adult elective cranial neurosurgery, reflecting persistent uncertainty in balancing thromboembolic and haemorrhagic risk. Quantitative analysis indicates that much of this variation represents true clinical equipoise and this has direct implications for evidence generation [32,33], identifying a clear opportunity for pragmatic randomised controlled trials. Such trials could address key unresolved questions, including the optimal timing of post-operative chemoprophylaxis and whether imaging-gated initiation improves safety without compromising effectiveness.

By mapping contemporary real-world practice across diverse health-system contexts, this study provides a necessary empirical foundation for rational trial design and future guideline development. In the absence of evidence generated through such trials, further guideline expansion alone is unlikely to reduce practice variation, and decision-making will remain driven by individual risk tolerance rather than comparative effectiveness data. Addressing this evidence gap is essential to developing harmonised, globally relevant strategies that improve the safety and consistency of VTE prevention in cranial neurosurgery.

## Data Availability

Raw survey data is available on reasonable request providing institutional permissions are met

